# Infection Risk Shifts of Protests During Pandemics

**DOI:** 10.1101/2023.04.15.23288618

**Authors:** Lennart Kraft, Michael Niekamp

## Abstract

This research article examines the dual impact of protests on COVID-19 spread, a challenge for policymakers balancing public health and the right to assemble. Using a game theoretical model, it shows that protests can shift infection risks between counties, creating a dilemma for regulators. The empirical study analyzes two German protests in November 2020 using proprietary data from a bus-shuttle service, finding evidence to support the assumption that protests can shift infection risks. The article concludes by discussing the implications of these findings for policymakers, highlighting that regulators’ individually rational strategic decisions may lead to inefficient outcomes.

## 1 Introduction

To mitigate the spread of diseases, particularly COVID-19, policymakers implement non-pharmaceutical interventions such as social distancing, restrictions on gatherings, or lockdowns. Yet, parts of the population disagree with those interventions and participate in protests. Regulators are concerned about such protests because they can reinforce the spread of diseases. However, prohibiting protests might violate protesters’ fundamental right to assemble. Since regulators must solve this fundamental conflict of interests, they need to know about a protest’s epidemic impact.

This article aims to provide information on protests’ epidemic impact by examining protests’ dual effect on the COVID-19 infection rate in protesters’ places of residence and the protests’ locations. An accumulation of citizens in a location may lead to an increase in the location’s COVID-19 infection rate if they do not comply with measures that mitigate the spread of COVID-19 (e.g., social distancing and mask-wearing Kwon et al. (2021), Wellenius et al. (2021)). Since an accumulation of citizens in one location decreases the number of citizens in another location, the other location’s COVID-19 infection rate might decrease. Thus, protests can shift the infection rate induced by protesters between locations – from protesters’ places of residence to protest locations – mitigating the spread in the former and reinforcing the spread in the later locations. In addition, after protests, returning protesters might have become infected and carry the infection back to their places of residence, mitigating the infection risk decrease. Such opposed impacts, however, can constitute a dilemma for regulators. Specifically, if one county’s citizens face the infection risk from their county’s and other counties’ protesters, each county might decide to prohibit protests. However, a decrease in the COVID-19 infection rate in protesters’ places of residence may (over-)compensate for an increase in the protest’s location. Thus, the average citizen might benefit if at least one county allowed a protest.

This article derives this conflict in a game theoretical model in which protests can shift infection risks between counties and tests the assumption that infection risk shifts can occur in an empirical study. In testing this assumption, it examines two protests in Germany happening in November 2020 to estimate protests’ dual effects on protesters’ places of residence and protests’ location, capturing the infection risk shift. The empirical study builds upon proprietary data of a bus-shuttle service that brought protesters from different German counties to both protests. Access to the ZIP codes of protesters using the bus-shuttle service and the number of COVID-19 infections per county enables the identification of the protest’s impact on the growth rates of COVID-19 infections in protests’ locations and protesters’ places of residence.

The remainder of the article is structured as follows. Section 2 provides background information on the protests examined in this article and outlines insights from previous literature on protests’ impact on the spread of COVID-19. Section 3 presents regulators’ dilemma based on a game theoretical model. Subsequently, Section 4 describes the data, outlines the methodological approach, and presents the results. Finally, Section 5 summarizes and concludes.

## 2 Insights on Protests’ Epidemic Impact

### 2.1 Summary of Background Information

The protests examined in this article took place in Leipzig and Berlin, Germany, on November 7, 2020, and November 18, 2020. Specifically, the German initiative “Querdenken” invited citizens to protest against policies aimed at mitigating the spread of COVID-19. According to official reports, at least 20,000 protesters participated in the protest in Leipzig. While the regulating authority required protesters to comply with some mitigation policies (e.g., social distancing and wearing masks), many protesters did not comply. Therefore, the police declared the protest dissolved but could not prevail with it (Deutschlandfunk 2020). Similarly, about 8,000 protesters gathered in Berlin on November 18, 2020, to protest against regulators’ non-pharmaceutical interventions. Like before, many protesters did not comply with the requirements of protests, and the police even had to use water guns to end the protest (Rundfunk Berlin-Brandenburg 2020).

These protests may differ from other gatherings where participants comply with required protective measures (e.g., social distancing and wearing masks). Hence, we note that the findings of this article may be specific to protests whose participants do not comply with protective measures, as also suggested by previous literature, summarized below.

### 2.2 Insights from Previous Literature

Insights from previous literature indicated that travel restrictions, social distancing, and mask-wearing reduced the spread of COVID-19 (Kwon et al. (2021), Wellenius et al., (2021) Ledesma et al. (2022)) and that mask-wearing mitigated social-distancing efforts, thus, mitigating the reduction in the spread of COVID-19 (Liebst et al. 2022). Since many protesters in Leipzig and Berlin did not comply with such mitigation strategies, we expect that these protests increased the spread of COVID-19 in these locations.

Concerning the consequences of gatherings, Dave et al. (2021) reported a positive impact of a gathering of motorcycle fans without a mitigation strategy on the spread of COVID-19 in the US. In contrast, Dave et al. (2020) reported that “Black Lives Matter” gatherings, where mitigation strategies were effective, did not affect the spread of COVID-19. Neyman et al. (2021) also analyzed “Black Lives Matter” Gatherings and found that infection rates in counties that participated in these gatherings were larger than in the other counties. Fischer(2022) reported that football matches increased the number of infections by three to seven percent and no moderating effect of stricter hygiene restrictions. However, Fischer(2022) identifies local mobility as underlying mechanism, suggesting that outdoor mass gatherings can increase infections. Similarly, Donsimoni et al. (2020) model the number of COVID-19 infections in Germany and found that interventions that lower the contact rate between individuals decrease the number of COVID-19 infections.

These findings are mostly consistent with previous literature outlining the protective effects of mitigation strategies. Similarly to these studies, we estimate the impact of mass gatherings on the spread of COVID-19 where protests occur. Additionally, this article examines the impact of mass gatherings on the spread of COVID-19 in participants’ places of residence, highlighting the dual role of protests in different locations.

## 3 Presentation of Regulators’ Dilemma

Regulators across counties need to decide whether to allow or prohibit protests. The possibility to exercise the right to assemble is valuable per se. However, our article focuses on the infection risk shifts to outline the conditions under which citizens might benefit from protests, even when neglecting the utility they might gain from this right. Below, we outline the game theoretical model that captures the core idea: protests might increase the spread of COVID-19 in the protest’s location, decrease it in the protesters’ places of residence, and the latter effect might (over-)compensate for the former. However, since each county regulator has an incentive not to allow a protest in its county, no protest might occur, and the equilibrium outcome could be inefficient (i.e., not maximizing citizens’ utility, respectively, not minimizing the infection risk).

### 3.1 Overview of Model

Table 1 provides an overview of the model. The model consists of two counties (X and Y). The regulators of each county select a strategy that determines whether a protest happens in their county (Protest) or not (No Protest). The combination of strategies leads to four different cells that represent different outcomes (i.e., different numbers of infected citizens) for X (upper part of a cell) and Y (lower part of a cell). For example, the strategy set (No Protest & No Protest) leads to a number of infected citizens equal to *N*_*x*_ *· r*_*b,x*_ for X and *N*_*y*_ *· r*_*b,y*_ for Y. Specifically, the number of infected citizens results from different parameters, explained below:

- *N*_*x*_ (*N*_*y*_) represent the number of citizens in *X* (*Y*).
- *r*_*b,x*_ (*r*_*b,y*_) represent the baseline infection rate in *X* (*Y*).
- *r*_*p,x*_ (*r*_*p,y*_) represent the increase in infection rates in *X* (*Y*) if a protest happens in *X* (*Y*) but not in *Y* (*X*).
- *r*_*np,x*_ (*r*_*np,y*_) represent the decrease in infection rates in *X* (*Y*) if a protest happens in *Y* (*X*) but not in *X* (*Y*).
- *r*_*p,xy*_ represents the increase in infection rates in *X* and *Y* if a protest happens in *X* and *Y*.

**Table 1:** Description of Regulators’ Strategies and Resulting Number of Infected Citizens

|  |  | Regulator Y |  |
| --- | --- | --- | --- |
|  |  | No Protest | Protest |
| Regulator X | No Protest | $N_x \cdot r_{b,x};$<br>$N_y \cdot r_{b,y}$ | $N_x \cdot (r_{b,x} - r_{np,x});$<br>$N_y \cdot (r_{b,y} + r_{p,y})$ |
| | Protest | $N_x \cdot (r_{b,x} + r_{p,x});$<br>$N_y \cdot (r_{b,y} - r_{np,y})$ | $N_x \cdot (r_{b,x} + r_{p,xy});$<br>$N_y \cdot (r_{b,y} + r_{p,xy})$ |

### 3.2 Implications of Model

Irrespective of the specific parameter values, both regulators will select “No Protest” as their dominant strategy to minimize the number of infections in their county. However, a regulator that aims to minimize the total number of infections (i.e., in *X* and *Y*) may select a different strategy set. More specifically, the total number of infections will be lower if a protest occurs only in *X* (*Y*) if the number of citizens in *Y* (*X*) is sufficiently larger than in *X* (*Y*). The number of citizens in *Y* (*X*) is sufficiently larger if equation (1) (equation (2)) holds, suggesting that a protest may lower the total number of infections by shifting the infection risk of a county with many citizens to a county with fewer citizens.

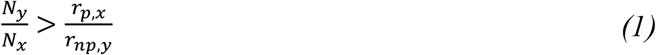

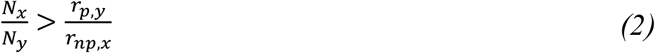

## 4 Presentation of Empirical Study

### 4.1 Description of Aim

The efficiency gain outlined in the previous section is possible because of the assumption that protests can shift infection risks between counties. Thus, the empirical study will test this assumption by examining whether infection risk shifts occurred between protesters’ places of residence and protest locations during protests and estimate the parameters *r*_*p,x*_ and *r*_*np,y*_ (or, respectively, *r*_*p,y*_ and *r*_*np,x*_). Specifically, it aims to compare the infection risk when there is no protest (i.e., the upper left cell in Table 1) to the infection risk if there is one protest (i.e., the lower left cell or the upper right cell in Table 1). For both counties, the study estimates the difference between these outcomes (i.e., (*r*_*b,x*_ + *r*_*p,x*_) − *r*_*b,x*_ = *r*_*p,x*_ for the protest county, defind as *X*, and (*r*_*b,y*_ − *r*_*np,y*_) − *r*_*b,y*_ = *r*_*p,y*_ for the county without a protest, defined as *Y*).

### 4.2 Overview of Datasets

Table 2 describes the two datasets analyzed in this study. The “Infection Dataset” provides each county’s official number of COVID-19 infections. The “Protest Dataset” provides the ZIP codes of protesters that used the data provider’s bus-shuttle service to travel to the protests in Leipzig and Berlin. We use the “Infection Dataset” to analyze the spread of COVID-19 in the protests’ location and combine it with the ZIP information of the “Protest Dataset” to examine the spread of COVID-19 in protesters’ places of residence.

**Table 2:** Overview of Datasets

| Name | Source | Number of counties | Time-Period | Description of Main Variables |
| --- | --- | --- | --- | --- |
| Infection Dataset | www.rki.de | 401 | September 12, 2020 – December 16, 2020 | Number of COVID-19 Infections |
| Protest Dataset | Kaden-Reisen | 401 | October 10, 2020 – December 16, 2020 | Protesters’ Places of Residence (ZIP Codes) |

Table 3 provides descriptive statistics of the datasets’ main variables across all 401 counties on November 7, 2020. The average cumulative number of COVID-19 infections equals 1,679, the average infection rate (i.e., defined as the cumulative number of COVID-19 infections divided by the number of citizens) equals 0.78%, and the average (weekly) growth rate of COVID-19 infections equals 22%. 64% and 60% of all counties are the places of residence of a protester participating in Leipzig’s or Berlin’s protest. On average, 2.60 and 2.05 protesters per county used the bus shuttle service to participate in the protest in Leipzig and Berlin. Thus, our data contains 1,044 and (=401·2.60) 823 (=401*2.05) protesters for Leipzig’s and Berlin’s protests. These values represent approximately 5.22% (=1,044/20,000) and 10.29% (=823/8,000) of the protesters estimated to have participated in these protests.

**Table 3:**
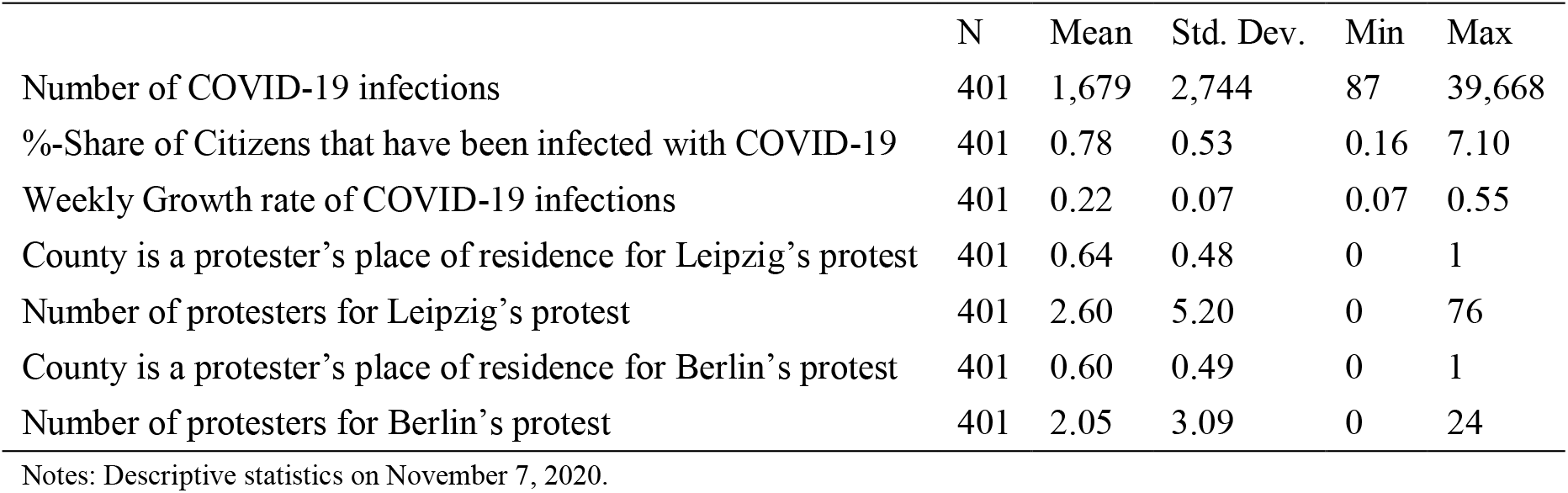
Descriptive Statistics of Datasets

### 4.3 Description of Methods

The empirical identification strategy builds upon Hsiang et al. (2020), who suggested comparing growth rates of COVID-19 infections instead of cumulative COVID-19 infections or COVID-19 infection rates. The first analysis compares such growth rates in protests’ locations (i.e., Leipzig and Berlin) before and after the protests (i.e., November 7, 2020, and November 18, 2020). The second analysis compares these growth rates in protesters’ places of residence before and after these protests.

Both analyses build upon the difference-in-differences methodology to control for time trends in the growth rates of COVID-19 infections by considering other counties as a control group. Each analysis uses the synthetic difference-in-differences methodology (Arkhangelsky et al. 2021) to create an artificial control group that satisfies parallel pre-treatment trends, a requirement to identify the protests’ impact on growth rates of COVID-19 infections via difference-in-differences approaches.

The control group for the Leipzig protest consists of counties that are neither Berlin, Leipzig, nor the place of residence for protesters using the shuttle service to participate in the protest in Leipzig. The control group for the Berlin protest consists of counties that are neither Berlin, Leipzig, nor the place of residence for protesters using the shuttle service to participate in the protest in Leipzig or Berlin.

Notably, it does not matter whether we identify the impact on growth rates of COVID-19 infections or infection rates (i.e., a county’s number of COVID-19 infections divided by its number of citizens) if the number of citizens is constant over the comparison period. Hence, the impacts identified in the analysis represent the parameters underlying the game theoretical model presented in Section 3. For example, a 3%p (percentage point) impact on the growth rate of COVID-19 infections represents a 3%p increase in the growth rates of infection rates and, thus, a 3% increase in infection rates.

### 4.4 Presentation of Results

Table 4 provides the estimates of protest’ impact on the growth rates of COVID-19 infections in Leipzig, Berlin, the places of residence of protesters, and their respective synthetic control groups. Model 1 shows that Leipzig’s weekly growth rates of COVID-19 infections increased by 3.67%p due to the protest (=exp(0.0360)-1). In contrast, the weekly growth rates in protesters’ places of residence decreased by 0.97%p (=exp(−0.0097)-1). Surprisingly, and in contrast to Leipzig, we find that Berlin’s weekly growth rates decreased after its protest by 4.88%p (=exp(0.0476)-1). For protesters’ places of residence, we again find decreases in their weekly growth rates by 0.42%p (=exp(−0.0042)-1), similar to the impact of Leipzig’s protest in protesters’ places of residence.

**Table 4:** Estimation Results for Protests’ Impact on Growth Rates of COVID-19 Infections

| Model | 1 | 2 | 3 | 4 |
| --- | --- | --- | --- | --- |
| Dependent Variable | Growth Rate of COVID-19 Infections | Growth Rate of COVID-19 Infections | Growth Rate of COVID-19 Infections | Growth Rate of COVID-19 Infections |
| Protest | Leipzig | Leipzig | Berlin | Berlin |
| Perspective | Protest's Location | Protesters' Places of Residence | Protest's Location | Protesters' Places of Residence |
| Main Variables |  |  |  |  |
| Protest | 0.0360<br>(0.85) | -0.0097*<br>(-1.90) | -0.0476<br>(-1.01) | -0.0042<br>(-0.41) |
| Control Variables |  |  |  |  |
| County FE | Yes | Yes | Yes | Yes |
| Time Trends | Yes | Yes | Yes | Yes |
| Model Statistics |  |  |  |  |
| Number of Treatment Counties | 1 | 256 | 1 | 39 |
| Number of Control Counties | 142 | 142 | 103 | 103 |
| Total Number of Counties | 143 | 398 | 104 | 142 |
| Number of Days | 84 | 84 | 84 | 84 |
| Total Number of Observations | 12,012 | 33,432 | 8,736 | 11,928 |

Notes: \* $p < 0.10$ . Estimations are based on Stata's script (i.e., "sdid") to implement the synthetic difference-in-differences approach (Arkhangelsky et al. 2021).

Figure 1 illustrates these results by comparing the growth rates of counties where the protest occurred (see top left and right panel) and where protesters came from (see bottom left and right panel) with the respective synthetic control groups. Each panel shows that the synthetic difference-in-differences methodology successfully implemented parallel pre-treatment trends. Moreover, each panel shows the different growth rate paths between these counties, outlining the impact development over time.

**Figure 1:**
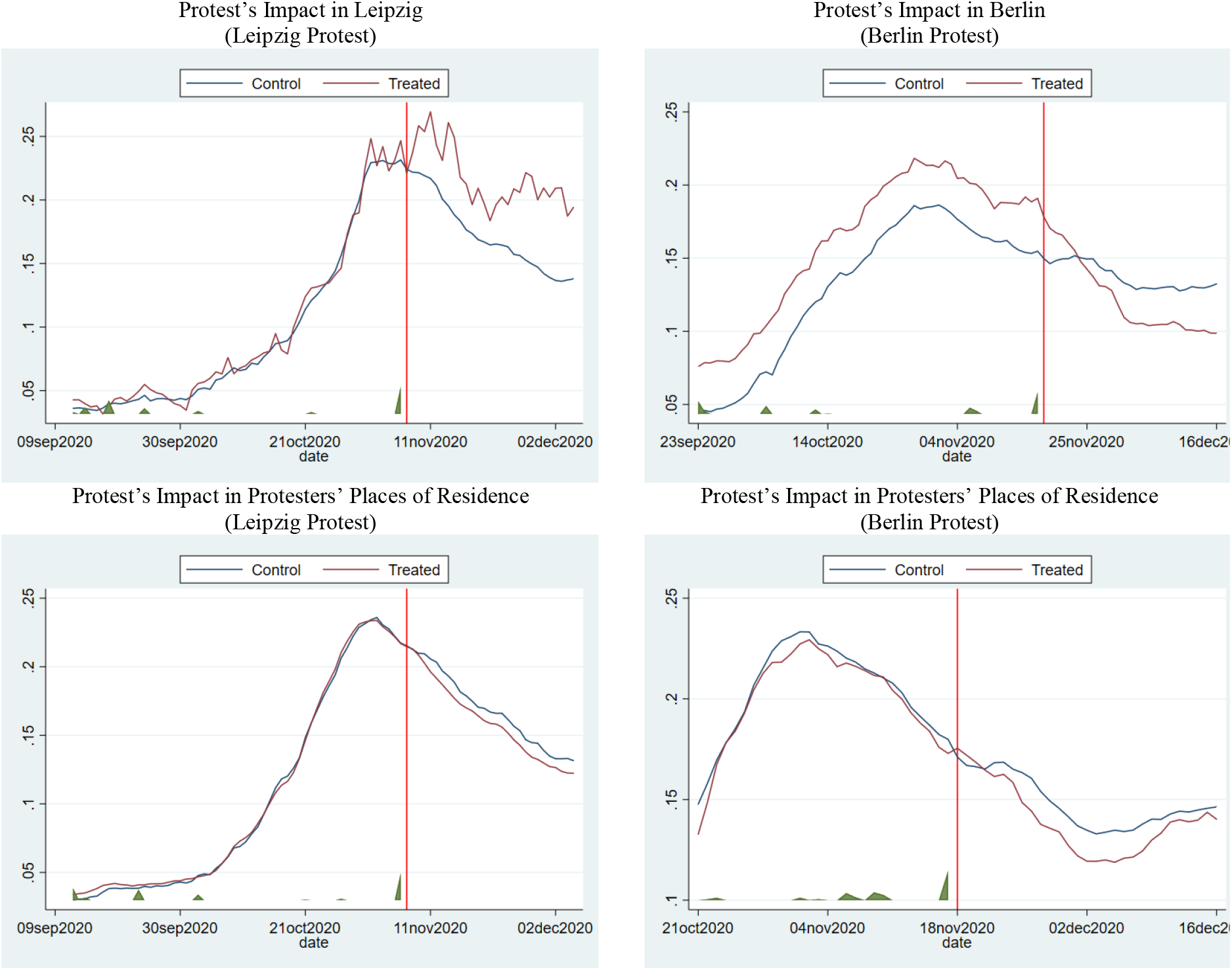
Illustration of Growth Rates of COVID-19 Infections Notes: Graphs are based on Stata’s script (i.e., “sdid”) to implement the synthetic difference-in-differences approach (Arkhangelsky et al. 2021). Red lines refer to weekly growth rates in COVID-19 infections of treated counties. Blue lines refer to weekly growth rates in COVID-19 infections of the synthetic control groups

### 4.5 Discussion of Results

The 0.97%p and 0.42%p reductions in growth rates of COVID-19 infections in protesters’ places of residence are consistent with the hypothesis that protests may shift the infection risk induced by protesters from their places of residence to where protests occur. We find mixed evidence of protests’ impact on growth rates of COVID-19 infections in locations where protests occur. Both estimates are statistically insignificant and also differ in their sign. The statistical insignificance may indicate no substantial impact but may also result from a lack of statistical power since only one county was available for the treatment group. Below, we outline potential reasons to explain our findings, starting with the results of protests’ epidemic impact on protests’ location, followed by their epidemic impact on protesters’ places of residence.

Infection rates in Leipzig and Berlin might have increased because protesters did not comply with the required protective measures and infected residents with COVID-19, or Leipzig and Berlin’s citizens conducted more COVID-19 tests. In addition, Berlin’s infection rate might have decreased because Berlin’s residents were aware of the epidemic impact of Leipzig’s protest. Thus, they might have increased their effort to protect themselves from becoming infected, outweighing the additional infection risk induced by protesters.

Infection rates in protesters’ places of residence might have decreased because protesters became infected but did not test themselves once they returned home. So, actual infection rates in protesters’ places of residence might have been higher. However, such behavior must have been different before the protests to bias the estimated effect because the models control for time trends. Alternatively, protesters might have returned home being infected with COVID-19 but isolated themselves and gained immunity afterward, decreasing infection rates in their places of residence. Alternatively, other citizens living in protesters’ places of residence might have also increased their effort to protect themselves from becoming infected in response to the protests. Ultimately, it remains unclear which exact mechanism could explain protests opposed impacts on the growth rates of COVID-19 infection in these locations.

## 5 Summary and Conclusion

Citizens’ fundamental right to assemble can conflict with their and others’ rights to be protected from the potential harm caused by exercising this right. This conflict became evident during the COVID-19 pandemic when citizens participated in protests and violated regulatory requirements of these protests, such as mask-wearing or social distancing. Consequently, such protests induce a fundamental conflict of interest for regulators. On the one hand, regulators should allow protests to let citizens exercise their fundamental right to assemble. On the other hand, regulators should prohibit protests to protect citizens from becoming infected because of such protests.

This article aimed to provide empirical insights into the epidemic impact of protests to inform regulators about the consequences of allowing protests. Specifically, it examined the dual effect of protests on the growth rate of COVID-19 infections in two location types. It finds that protests can reduce these growth rates in protesters’ places of residence by up to 0.97%p (p<0.10) but may increase them in the locations where protests occur by up to 3.67%p (p>0.10), potentially shifting the infection risk from the former locations to the latter.

Therefore, we conclude that allowing protests may increase COVID-19 infections where protests take place. However, allowing protests could also benefit citizens’ right to assemble and reduce COVID-19 infections in locations where protesters come from. So, our results suggest that protests could have opposed epidemic impacts. These opposing impacts result in a dilemma because regulators’ individually rational strategic decisions may not lead to an efficient equilibrium that minimizes the infection risk of all citizens.

## Data Availability

The datasets are proprietary. Interested researchers can contact one of the authors via email to discuss access to the data.

## 8 Author Contributions

LK and MN conceptualized the research idea and conducted literature research. MN collected the data. LK designed empirical methods and analyzed the data. LK and MN interpreted the results and wrote the article. LK and MN reviewed and revised the article.

## 10 Additional Information

Correspondence and requests for materials should be addressed to Lennart Kraft

## References

Arkhangelsky, Dmitry, Susan Athey, David A. Hirshberg, Guido W. Imbens, and Stefan Wager. 2021. “Synthetic Difference-in-Differences.” American Economic Review 111 (12): 4088–4118. https://doi.org/10.1257/aer.20190159.

Dave, Dhaval M., Andrew I. Friedson, Kyutaro Matsuzawa, Joseph J Sabia, and Samuel Safford. 2020. “Black Lives Matter Protests, Social Distancing, and Covid-19.” NBER Working Papers, no. 27408.

Dave, Dhaval, Drew McNichols, and Joseph J. Sabia. 2021. “The Contagion Externality of a Superspreading Event: The Sturgis Motorcycle Rally and Covid-19.” Southern Economic Journal 87 (3): 769–807. https://doi.org/10.1002/soej.12475.

Deutschlandfunk. 2020. “‘Querdenken’-Bewegung -Kritik und Konsequenzen nach Leipziger Demonstrationen.” Deutschlandfunk. November 9, 2020. https://www.deutschlandfunk.de/querdenken-bewegung-kritik-und-konsequenzen-nach-leipziger-100.html.

Donsimoni, Jean Roch, René Glawion, Bodo Plachter, and Klaus Wälde. 2020. “Projecting the Spread of COVID-19 for Germany.” German Economic Review 21 (2): 181–216. https://doi.org/10.1515/ger-2020-0031.

Fischer, Kai. 2022. “Thinning out Spectators: Did Football Matches Contribute to the Second COVID-19 Wave in Germany?” German Economic Review 23 (4): 595–640. https://doi.org/10.1515/ger-2021-0060.

Hsiang, Solomon, Daniel Allen, Sébastien Annan-Phan, Kendon Bell, Ian Bolliger, Trinetta Chong, Hannah Druckenmiller, et al. 2020. “The Effect of Large-Scale Anti-Contagion Policies on the Covid-19 Pandemic.” Nature 584 (7820): 262–67. https://doi.org/10.1038/s41586-020-2404-8.

Kwon, Sohee, Amit D. Joshi, Chun-Han Lo, David A. Drew, Long H. Nguyen, Chuan-Guo Guo, Wenjie Ma, et al. 2021. “Association of Social Distancing and Face Mask Use with Risk of COVID-19.” Nature Communications 12 (1): 3737. https://doi.org/10.1038/s41467-021-24115-7.

Ledesma, Jorge R., Lin Zou, Stavroula A. Chrysanthopoulou, Danielle Giovenco, Aditya S. Khanna, and Mark N. Lurie. 2022. “Community Mitigation Strategies, Mobility, and COVID-19 Incidence Across Three Waves in the United States in 2020.” Epidemiology Publish Ahead of Print (September). https://doi.org/10.1097/EDE.0000000000001553.

Liebst, Lasse S., Peter Ejbye-Ernst, Marijn de Bruin, Josephine Thomas, and Marie R. Lindegaard. 2022. “No Evidence That Mask-Wearing in Public Places Elicits Risk Compensation Behavior During the COVID-19 Pandemic.” Scientific Reports 12 (1): 1511. https://doi.org/10.1038/s41598-022-05270-3.

Neyman, Greg, and William Dalsey. 2021. “Erratum: JPH-20-1234-Black Lives Matter Protests and COVID-19 Cases: Relationship in Two Databases.” Journal of Public Health 43 (2): 225–27. https://doi.org/10.1093/pubmed/fdab053.

Rundfunk Berlin-Brandenburg. 2020. “Polizei beendet Demo mit Wasserwerfern -365 Festnahmen.” November 18, 2020. https://www.rbb24.de/politik/thema/2020/coronavirus/beitraege_neu/2020/11/demonst rationen-corona-gegner-bundestag-infektionsschutzgesetz.html.

Wellenius, Gregory A., Swapnil Vispute, Valeria Espinosa, Alex Fabrikant, Thomas C. Tsai, Jonathan Hennessy, Andrew Dai, et al. 2021. “Impacts of Social Distancing Policies on Mobility and COVID-19 Case Growth in the US.” Nature Communications 12 (1): 3118. https://doi.org/10.1038/s41467-021-23404-5.

